# A multidimensional analysis reveals distinct immune phenotypes and the composition of immune aggregates in pediatric acute myeloid leukemia

**DOI:** 10.1101/2023.03.03.23286485

**Authors:** Joost B. Koedijk, Inge van der Werf, Livius Penter, Marijn A. Vermeulen, Farnaz Barneh, Alicia Perzolli, Joyce I. Meesters-Ensing, Dennis S. Metselaar, Thanasis Margaritis, Marta Fiocco, Hester A. de Groot-Kruseman, Rubina Moeniralam, Kristina Bang Christensen, Billie Porter, Kathleen Pfaff, Jacqueline S. Garcia, Scott J. Rodig, Catherine J. Wu, Henrik Hasle, Stefan Nierkens, Mirjam E. Belderbos, C. Michel Zwaan, Olaf Heidenreich

**Author notes:** Corresponding author and lead contact: Olaf Heidenreich, Princess Máxima Center for Pediatric Oncology, 3584 CS Utrecht, The Netherlands. Equal contribution (C. Michel Zwaan, Olaf Heidenreich).

## Abstract

Because of the low mutational burden and consequently, fewer potential neoantigens, children with acute myeloid leukemia (AML) are thought to have a T cell-depleted or ‘cold’ tumor microenvironment and may have a low likelihood of response to T cell-directed immunotherapies. Understanding the composition, phenotype, and spatial organization of T cells and other microenvironmental populations in the pediatric AML bone marrow (BM) is essential for informing future immunotherapeutic trials about targetable immune-evasion mechanisms specific to pediatric AML. Here, we conducted a multidimensional analysis of the tumor immune microenvironment in pediatric AML and non-leukemic controls. We demonstrated that nearly one-third of pediatric AML cases has an immune-infiltrated BM, which is characterized by a decreased ratio of M2-to M1-like macrophages. Furthermore, we detected the presence of large T cell networks, both with and without colocalizing B cells, in the BM and dissected the cellular composition of T- and B cell-rich aggregates using spatial transcriptomics. These analyses revealed that these aggregates are hotspots of CD8^+^ T cells, memory B cells, plasma cells and/or plasmablasts, and M1-like macrophages. Collectively, our study provides a multidimensional characterization of the BM immune microenvironment in pediatric AML and indicates starting points for further investigations into immunomodulatory mechanisms in this devastating disease.

## Introduction

The abundance and phenotype of intratumoral T cells are crucial for the effectiveness of T cell-directed immunotherapies such as immune checkpoint inhibitors (ICIs) and bispecific T cell-engagers^1–4^. Because of the low mutational burden and consequently, fewer potential neoantigens, children with acute myeloid leukemia (AML) are thought to have a T cell-depleted or ‘cold’ tumor microenvironment and therefore may have a low likelihood of response to ICIs and bispecific T cell-engagers^5–8^. Although clinical trials in adult AML patients treated with ICIs and bispecific T cell-engagers, whether as mono- or combination therapy, have largely been unsuccessful, a small subset of patients showed exceptional responses^4, 9–15^. This suggests that there may be specific subgroups that could benefit from these T cell-directed immunotherapies. To pave the way for successful ICI- and bispecific T cell-engager immunotherapies in both adult and pediatric AML, a better understanding of the heterogenous landscape of bone marrow (BM)-infiltrating T cells and the surrounding tumor microenvironment is needed. While recent studies have provided insights on this matter in adult AML, such as the identification of a relatively low presence of exhausted CD8^+^ T cells in the tumor microenvironment compared to cancers that respond well to ICIs^16, 17^, there is a paucity of data on the BM immune microenvironment in pediatric AML^11, 18–20^. In addition to the need for a quantitative and qualitative assessment of the pediatric AML BM immune microenvironment, emerging research in solid cancers has shown that the spatial organization of the immune response is highly relevant for ICI efficacy^21^. For instance, ‘excluded’ tumors, where the immune response cannot invade the tumor bed, show poor responses to ICI^22^. Furthermore, the presence of intratumoral immune aggregates, such as tertiary lymphoid structures (TLS), is associated with improved ICI responses in many solid cancers^23–28^. Despite its potential importance, the spatial organization of the immune response in hematological cancers remains understudied. Therefore, we generated a multidimensional view of the tumor immune microenvironment in treatment-naïve *de novo* pediatric AML, to inform future immunotherapeutic trials about potentially targetable immune-evasion mechanisms specific to this patient population. We identified distinct BM immune phenotypes and dissected the composition of immune aggregates in the BM of pediatric AML, which encourage further investigations into immunomodulatory mechanisms in this devastating disease.

## Materials/Subjects and Methods

### Ethical regulation

This study complies with all relevant ethical regulations and was approved by the Institutional Review Boards of the Princess Máxima Center for Pediatric Oncology (PMCLAB2021.207 & PMCLAB2021.238), the Scientific Committee of the Dutch Nationwide Pathology Databank (PALGA: lzv2021-82)^29^ and at participating sites of the ETCTN/CTEP 10026 study ^12, 13^. All patients treated at the Princess Máxima Center, Aarhus University Hospital, and Dana-Farber Cancer Institute provided written consent for banking and research use of these specimens, according to the Declaration of Helsinki.

### Human patient samples

FFPE BM biopsies taken from the crista of children with treatment-naïve *de novo* AML and non-leukemic controls (Supplementary Methods) were obtained from the Princess Máxima Center Biobank (n=15), biobanks of 10 other Dutch hospitals (n=28; mediated by the Dutch National Tissue Portal) and the biobank of Aarhus University Hospital (n=29). BM biopsies of adult AML cases treated on the ETCTN/CTEP 10026 study (n=6) were collected at Dana-Farber Cancer Institute^13, 17^. Details on other pediatric and adult AML datasets, immunohistochemistry/immunofluorescence, digital image analysis, spatial transcriptomics, and immune-related gene expression profiling are provided in the Supplementary Methods.

### Statistical analyses

Statistical analyses were performed with GraphPad Prism V.9.3.0 (GraphPad Software, LA Jolla, CA, USA). Details on the various statistical tests used are provided in the Supplementary Methods. For spatial transcriptomics data, *P*<0.01 was considered statistically significant to correct for measuring multiple regions from the same biopsy. For all other comparisons, *P*<0.05 was considered statistically significant.

## Results

### A subset of pediatric AML patients has high T cell infiltration in the bone marrow

To determine whether the BM microenvironment in pediatric AML is characterized by high (‘hot’) or low (‘cold’) T cell infiltration, we performed immunohistochemistry (IHC) with antibodies against CD3 and CD8 on 82 formalin-fixed and paraffin-embedded (FFPE) BM biopsies from pediatric patients with treatment-naïve *de novo* AML (n=72), and age- and sex-matched non-leukemic individuals (n=10; **Fig. 1A**; representative images shown in **Fig. 1B-C**; collectively referred to as ‘primary study cohort’). Patient characteristics are depicted in **Table S1**. We found a trend towards a decreased abundance of the number of T cells and a significant decrease in CD8^+^ T cells in pediatric AML cases in comparison to non-leukemic controls (*P*=0.11 and *P*=0.011, respectively; **Fig. 1D-E**), similar to other observations in adult and pediatric AML^20, 31, 32^. The T- and CD8^+^ T cell infiltration ranged from 55 to 9832 cells/mm^2^ between individual pediatric AML cases, with a subset showing high T- and CD8^+^ T cell infiltration in the BM (above median of non-leukemic controls; n=22 and n=18, respectively; **Fig. 1D-E**).

**Figure 1.**
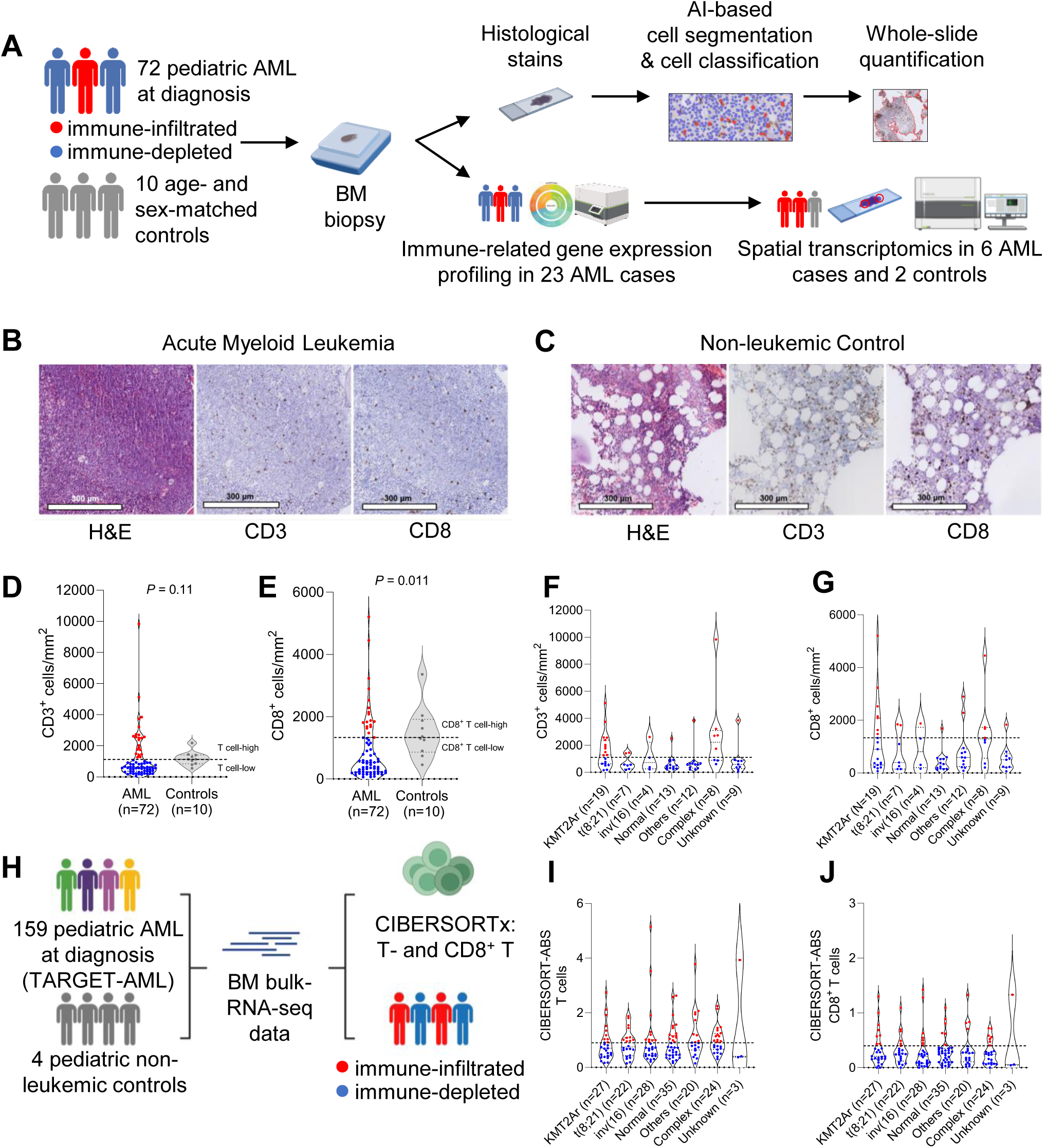
Characterizing T cell infiltration in pediatric AML cases and non-leukemic controls. (A) Schematic overview of the study population, used techniques, and the digital image analysis pipeline. AML cases are categorized in immune-infiltrated (red) and immune-depleted (blue) groups according to their T cell infiltration levels (above or below median of non-leukemic controls). (B-C) Representative bone marrow biopsy images from a treatment-naïve pediatric AML case (B) and a non-leukemic control (C) showing H&E staining, CD3^+^ T cells, and CD8^+^ T cells. White lobules indicate adipocytes. (D-E) Comparison of the normalized abundance of CD3^+^ T cells (D) and CD8^+^ T cells (E) in the bone marrow between pediatric AML cases and non-leukemic controls using the Mann-Whitney test. The dashed lines indicate the median CD3^+^ (D) and CD8^+^ (E) T cell abundance in non-leukemic controls, respectively. (F-G) Normalized abundance of CD3^+^ (F) and CD8^+^ T cells (G) per cytogenetic subgroup. ‘Normal’ indicates normal karyotype, while ‘Others’ is a merge of cytogenetic abnormalities different from the five defined cytogenetic subgroups. See Table S1. ‘Complex’ indicates cases with complex karyotype AML (≥3 chromosomal abnormalities). The dashed lines shown in Figure 1D-E are also shown in Figure 1F-G. (H) Schematic overview of the TARGET-AML cohort, the additional non-leukemic control group, the performed analysis (CIBERSORTx), and the subsequent categorization of patients into the immune-infiltrated or immune-depleted groups (based on median T- and CD8^+^ T cell abundance in non-leukemic controls). (I-J) Estimated absolute (ABS) abundance of T-(I) and CD8^+^ T (J) cells in the bone marrow of treatment-naïve pediatric AML cases in the TARGET-AML cohort. The dashed lines shown in Figure 1I-J indicate the estimated median bone marrow T- and CD8^+^ T cell abundance in four non-leukemic controls.

We next explored whether this heterogeneity in T cell infiltration reflected inherent differences in disease biology. Notably, eleven out of nineteen patients with *KMT2A*-rearranged AML (58%) and five out of eight patients with complex karyotype AML (63%) had high BM T cell infiltration (**Fig. 1F**). However, substantial heterogeneity in T cell infiltration was also present within both the *KMT2A*-rearranged and complex karyotype AML groups (**Fig. 1F-G**). Thus, both high and low levels of BM T cell infiltration were noted among *KMT2A*-rearranged and complex karyotype AML patients. Most cases in the other cytogenetic subgroups had low overall T- and CD8^+^ T cell infiltration (**Fig. 1F-G**). Specifically, twelve out of thirteen cases (92%) with normal karyotype AML had low total T- and CD8^+^ T cell infiltration. Among these thirteen cases, nine (69%) had both low T cell infiltration and a *FLT3*-ITD and/or *NPM1* mutation (**Table S1**), which have been associated with low T- and NK cell infiltration in adult AML^33^. Using diagnostic flow cytometry data of an independent cohort of 20 pediatric cases with normal karyotype AML (n=12 wildtype, n=8 *FLT3*-ITD and/or *NPM1* mutation), we confirmed this association (*P*=0.013; **Fig. S1A**).

Given the relatively small number of samples in some cytogenetic subgroups, we also reanalyzed a large bulk RNA-sequencing dataset of treatment-naïve pediatric AML BM aspirates (TARGET-AML cohort, n=159 patients; **Fig. 1H**)^34^. Using the immune deconvolution tool CIBERSORTx^17, 35, 36^, we estimated the absolute abundance scores of T- and CD8^+^ T cells, which ranged from 0.15 to 5.15 and 0 to 1.43, respectively (**Fig. 1H-J**). We categorized patients into overall T- and CD8^+^ T cell-high and -low groups based on the median estimated BM T- and CD8^+^ T cell abundance in four non-leukemic pediatric controls (BM bulk RNA-sequencing data^37^). The TARGET-AML cohort showed substantial heterogeneity in overall T- and CD8^+^ T cell levels across all cytogenetic subgroups, with only nine out of 27 *KMT2A*-rearranged AML cases (33%) having high T cell infiltration (**Fig. 1I-J**). Consequently, we hypothesized that specific *KMT2A*-rearrangements may be associated with different levels of T cell infiltration in the BM. We therefore compared the levels of T- and CD8^+^ T cells among cases with *KMT2A*-rearrangements with at least three samples per group (TARGET-AML cohort). This analysis suggested a lower abundance of CD8^+^ T cells in *KMT2A*::*ELL* AML compared to *KMT2A*::*MLLT3* AML (*P=*0.044; **Fig. S1B-C**), although the small sample size precluded firm conclusions. Among cases with complex karyotype AML, thirteen out of 24 cases (54%) had high T cell infiltration (**Figure 1I**). However, we were not able to identify any commonalities between those with high or low T cell infiltration. Other clinical factors including the abundance of leukemic blasts and AML differentiation stage showed no or only marginal correlation with T- and CD8^+^ T cell infiltration (**Fig. S1D-K)**.

To further characterize the immune profiles of pediatric AML cases with a high abundance of T cells in the BM, we categorized AML cases in the primary study cohort into two groups using the non-leukemic controls’ median CD3^+^ T cell abundance as the cut-off value (1133 cells/mm^2^; **Fig. 2A**). Focused IHC analyses of CD20 expression showed that patients in the CD3^+^ T cell-high group also had a significantly higher abundance of CD20^+^ B cells in comparison to the CD3^+^ T cell-low group (*P*<0.001; **Fig. 2B**). Accordingly, we termed the two groups ‘immune-infiltrated’ and ‘immune-depleted’ (n=22 and n=50, respectively). We subsequently investigated whether these groups demonstrated differences in clinical outcomes upon standard chemotherapy treatment (Supplementary Methods), as seen in many other cancer types^1^. In our cohort, we did not detect differences in overall survival (OS) between the immune-infiltrated and immune-depleted group (**Fig. S2A**). To increase statistical power, we repeated the analysis in the TARGET-AML cohort. Again, we did not detect differences in OS between the immune-infiltrated (CD3^+^ T cell-high, n=66) and immune-depleted (CD3^+^ T cell-low, n=93) groups (**Fig. S2B**), together suggesting that the extent of T cell infiltration in the BM at diagnosis is not critical for patient survival in pediatric AML cases treated with standard chemotherapy regimens. Taken together, we identified wide heterogeneity in T cell infiltration in the BM of pediatric AML with nearly one-third of cases (31%) showing notably high T cell infiltration. In addition, our findings suggest that specific genetic alterations may be linked to the extent of T cell infiltration in the BM of pediatric AML, although the exact underlying mechanisms remain to be unraveled^38, 39^.

**Figure 2.**
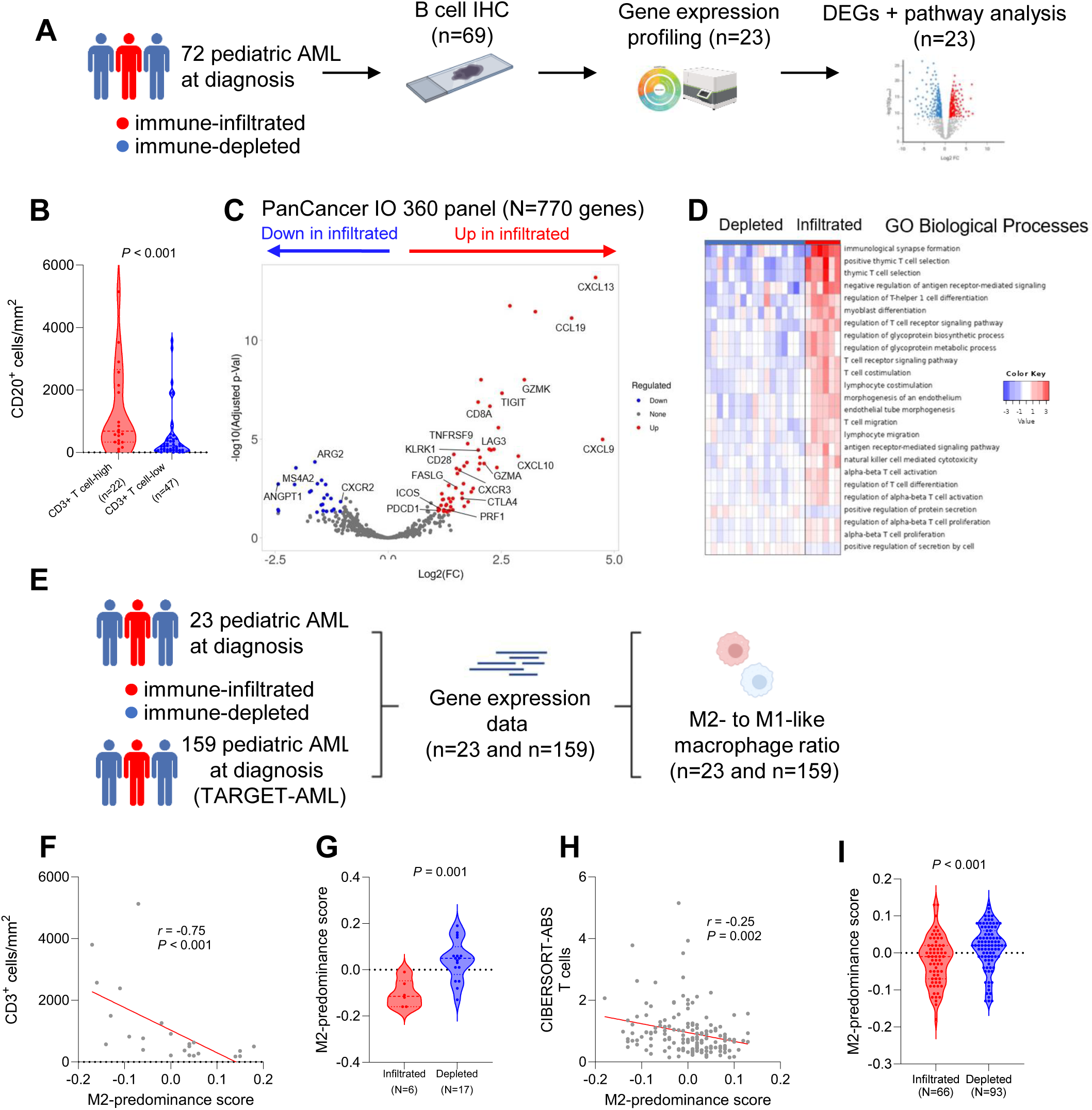
Transcriptional differences between immune-infiltrated and immune-depleted pediatric AML. (A) Schematic overview of the study population, used techniques, and analyses performed in Figure 2B-D. (B) Comparison of the normalized abundance of CD20^+^ B cells in the bone marrow of pediatric AML (CD20 stains available for 69 cases) with CD3^+^ T cell levels above or below the median of non-leukemic controls (later referred to as immune-infiltrated and immune-depleted, respectively; Mann-Whitney test). (C) Volcano plot of differentially expressed genes between immune-infiltrated (n=6) and immune-depleted (n=17) pediatric AML bone marrow biopsies, identified using *DEseq2* with an FDR cut-off of 0.05 and minimum fold change (FC) of 2. (D) Single-sample gene set enrichment analysis of differentially expressed genes between immune-infiltrated and immune-depleted cases using the GO Biological Processes gene set with an FDR cut-off of 0.05. WikiPathways-related results are shown in Table S2. (E) Schematic overview of the study populations for which gene-expression data was available (primary study cohort and TARGET-AML cohort), and the associated analysis. (F) Correlation plot of the negative correlation between the M2-predominance score and the normalized number of CD3^+^ T cells, calculated using Spearman correlation. (G) Comparison of the M2-predominance score between immune-infiltrated (n=6) and immune-depleted (n=17) cases using the Mann-Whitney test. (H) Correlation plot of the negative correlation between the M2-predominance score and the estimated abundance of T cells in the bone marrow of TARGET-AML cases, calculated using Spearman correlation. (I) Comparison of the M2-predominance score between immune-infiltrated (n=80) and immune-depleted (n=79) cases using the Mann-Whitney test.

### The ratio of M2- to M1-like macrophages is linked to the extent of T cell infiltration in the pediatric AML bone marrow

To elucidate mechanisms driving high- and low immune infiltration in the BM of pediatric AML, we examined the expression of immune-related genes in a cytogenetically representative subset of immune-infiltrated (n=6) and immune-depleted cases (n=17) using the NanoString PanCancer IO 360^TM^ panel (**Fig. 2A**). Using differential gene expression analysis, we identified genes related to T- and/or NK cells to be significantly upregulated in immune-infiltrated compared to immune-depleted pediatric AML (**Fig. 2C**), confirming our categorization of cases into these two groups. Regarding factors that either promote or restrict T cell infiltration, we found that immune-infiltrated cases demonstrated significantly higher expression of genes related to T cell-attracting chemokines (*CXCL9*, *CXCL10*), and their corresponding receptor (*CXCR3*; **Fig. 2C**). In line with this, pathway analysis using the GO biological processes and WikiPathways gene sets indicated that immune-infiltrated cases were enriched in T cell migration and chemokine signaling, suggesting that immune-depleted cases lack signals that attract T cells to the BM (**Fig. 2D**; full list of pathways in **Table S2**). In solid cancers, macrophages have been described as key players in attracting T cells^40^. Specifically, pro-inflammatory M1-like macrophages are known to be a primary source of T cell-attracting chemokines, while anti-inflammatory M2-like macrophages restrict T cell infiltration into the tumor^40, 41^. Previously, a gene expression-based score that reflects the ratio between M2- and M1-like macrophages (a score > 0 indicates M2-predominance, while a score < 0 indicates M1-predominance; collectively termed M2-predominance score) has been developed for five cancer types including AML^42^. We applied this score to our cohort and found that the M2-predominance score was negatively correlated with the BM T- and CD8^+^ T cell abundance (*r*=-0.75 [95% CI: −0.89 to −0.48], *P*<0.001; *r*=-0.50 [95% CI: - 0.76 to −0.12], *P*=0.014, respectively; **Fig. 2E,F** and **Fig. S2C**). Concordantly, we identified a significantly decreased M2-predominance score in immune-infiltrated compared to immune-depleted cases (*P*=0.001; **Fig. 2G**). Likewise, in the TARGET-AML cohort, we detected a (subtle) negative correlation between the M2-predominance score and the T- and CD8^+^ T cell abundance in the BM (*r*=-0.25 [95% CI: −0.39 to −0.10], *P*=0.002; *r=*-0.20 [95% CI: −0.35 to −0.05], *P*=0.011, respectively; **Fig. 2H** and **Fig. S2D**), and a significantly decreased M2-predominance score in the immune-infiltrated compared to the immune-depleted group (*P*<0.001.; **Fig. 2I**). Altogether, our data suggest that the M2-:M1-like macrophage ratio is linked to the extent of T cell infiltration in the BM of pediatric AML, in line with data from (preclinical) studies in other cancers^40, 42^.

### T cells cluster into aggregates in the bone marrow of pediatric AML

Several studies in solid cancers have identified the presence of immune aggregates in the tumor microenvironment, such as T- and B cell-rich structures resembling secondary lymphoid organs (TLSs) and T cell-rich structures lacking B cells^23–25, 43, 44^. These aggregates may represent sites of priming or re-activation of anti-tumor immune responses and have been associated with better responses to immune checkpoint inhibitors in many cancers types^23–28, 45, 46^. As TLSs and other immune aggregates often develop at sites of chronic inflammation^23–25^, we next asked whether similar structures were present in the BM of immune-infiltrated pediatric AML cases. Accordingly, we first investigated the number of T cell networks in pediatric immune-infiltrated (n=22), immune-depleted (n=50), and non-leukemic BM biopsies (n=10). In this analysis, T cell networks were defined as at least ten directly interacting T cells (≤10 μm between adjacent nuclei), to avoid classifying randomly dispersed T cells as networks (**Fig. 3A**)^44^. We found that T cell networks were present in both AML groups and controls, although they were significantly more frequent in the BM of immune-infiltrated compared to immune-depleted cases (*P*<0.001; **Fig. 3B** and **Table S1**; representative image of a T cell network shown in **Fig. 3C**). Moreover, controls had significantly more T cell networks in comparison to immune-depleted cases (*P*=0.027; **Fig. 3B**; representative images of immune-depleted biopsies shown in **Fig. S3A-B**).

**Figure 3.**
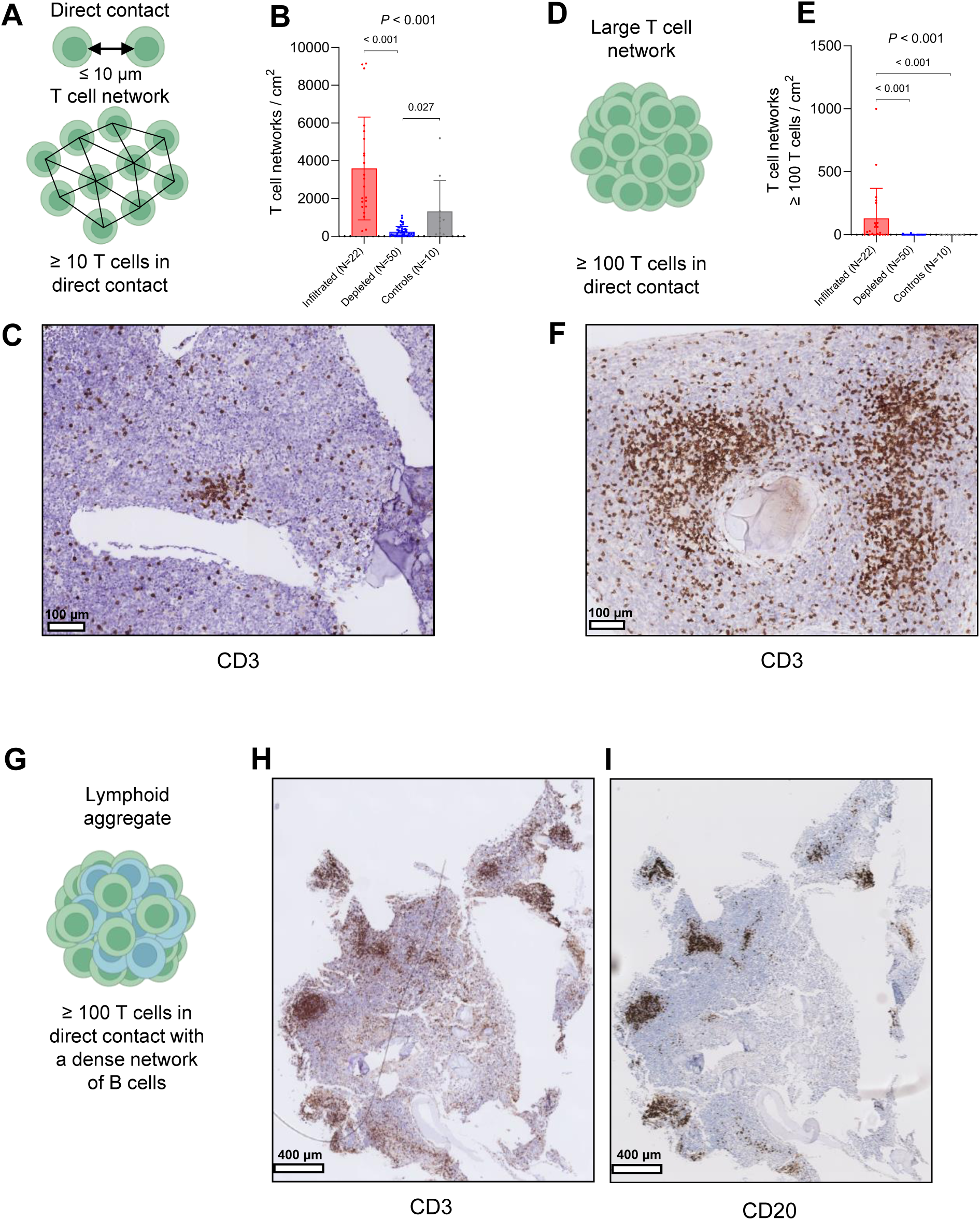
T cell networks in the bone marrow of pediatric AML. (A) Illustration of the identification of directly interacting T cells (above) and T cell networks (below) using Delaunay Triangulation. (B) Comparison of the normalized abundance of T cell networks between immune-infiltrated (n=22), immune-depleted (n=50), and non-leukemic control biopsies (n=10; Kruskal-Wallis followed by Dunn’s multiple comparisons test). (C) Representative image of a T cell network in a treatment-naïve pediatric AML patient. (D) Schematic of the definition of a large T cell network. (E) Comparison of the number of T cell networks with at least 100 T cells/network between immune-infiltrated (n=22), immune-depleted (n=50), and non-leukemic control biopsies (n=10; Kruskal-Wallis followed by Dunn’s multiple comparisons test). (G) Schematic of the definition of a lymphoid aggregate. (H-I) Representative images of lymphoid aggregates.

In addition, we identified large T cell networks, defined as at least 100 directly interacting T cells (**Fig. 3D**), to be more abundant in the BM of immune-infiltrated compared to immune-depleted cases (*P*<0.001; **Fig. 3E**; representative image shown in **Fig. 3F**). *KMT2A*-rearranged (9/17 cases; 53%) and complex karyotype AML (3/17 cases; 18%) were the most common cytogenetic subgroups among cases with large T cell networks (n=17 in total; **Table S1**). These large networks often colocalized with a dense network of B cells in immune-infiltrated cases (termed ‘lymphoid aggregates’; LA; **Fig. 3G**; 9/15 cases; 60%); representative images shown in **Fig. 3H-I, Fig. S3C-D**; 0/2 immune-depleted cases). However, the LAs identified in pediatric AML patients were not organized in a fashion seen in mature TLSs with an inner zone of B cells surrounded by T cells^23–25^. Instead, T- and B cells appeared to be mixed throughout the aggregate, as seen in immature TLSs and other immune aggregates (representative images shown in **Fig. 3H-I** and **Fig. S3C-D**)^25, 43^. In the BM of non-leukemic control biopsies, we did not identify LAs or large T cell-dominant networks with no or sparse B cells (representative image shown in **Fig. S3E**). Taken together, these data show that large networks of T cells, both with and without colocalizing B cells, are frequent in the BM of treatment-naïve immune-infiltrated pediatric AML cases.

Given that TLSs and other immune aggregates may function as sites of intratumoral immune priming that can lead to successful anti-tumor immunity in solid cancers^23–28, 45–48^, they may also serve as markers of leukemia-specific immune responses in hematological malignancies. Consequently, we performed pilot work to explore whether responses to immune checkpoint inhibitors were associated with the presence of LAs and/or large T cell-dominant networks in the AML BM. As data from pediatric AML cases treated with such therapies were not available, we applied multiplex immunofluorescence to pre- and post-treatment BM biopsies of transplant-naïve and post-transplant adult AML cases (three responders and three non-responders) treated with both ipilimumab and decitabine in the context of a clinical trial (NCT02890329^13, 17^; **Fig. S4A**). Accordingly, BM biopsies collected at baseline, time of best response, and end of treatment were stained with antibodies against CD3, CD20, and CD34 (n=17 in total; representative images of LAs and large T cell-dominant networks are shown in **Fig. S4B-D**; patient characteristics are depicted in **Table S3**). LAs were only observed in responders (response definitions in Supplementary Methods), although their time of appearance differed (**Fig. S4E-J**). Large T cell-dominant networks were present in two responders at baseline and at time of best response (AML1002 + AML1006; **Fig. S4E,G**). Among non-responders, one patient had large T cell-dominant networks at baseline (AML1010; **Fig. S4I**). Like pediatric AML, T- and B cells in lymphoid aggregates were not organized in distinct zones (**Fig. S4B-C**). Thus, in this exploratory analysis, we found that large T cell-dominant networks were present both at baseline and at time of best response in two out of three responders, whereas LAs were present in all three responders but at different time points. Although the patient heterogeneity and small cohort size preclude conclusions regarding the predictive utility of lymphoid aggregates and T cell dominant networks, our preliminary findings encourage future investigations into the association between immune aggregates and ICI response in both adult and pediatric AML, as done in solid cancers^26–28, 45^.

### Spatial transcriptomics unravels the composition of lymphoid aggregates in the bone marrow of pediatric AML

The identification of LAs prompted us to explore whether the profiles of these aggregates shared similarities with TLSs in solid cancers (i.e. several T cell subsets, germinal center B cells, plasma cells, dendritic cells)^23–25^. To accurately identify TLSs, several transcriptomic signatures have been proposed^25^: a 12-gene chemokine signature^49^ (‘12chem’; reflective of TLSs independent of their maturation stage), an 8-gene follicular helper T cell signature^50^ (‘Tfh’; reflective of mature TLSs), and a 29-gene TLS imprint signature^27^ (reflective of mature TLSs). We aimed to examine the expression of these signatures in both LA and BM regions without such aggregates. Towards this aim, we performed spatial transcriptomics using the GeoMx spatial transcriptomics platform^30^ on eight BM biopsies: four immune-infiltrated AML biopsies with LAs, and four reference biopsies (two immune-infiltrated AML biopsies without LAs and two non-leukemic control biopsies; patient characteristics are depicted in **Table S1**). Regions of interest (ROIs) included areas with LAs, ‘mixed’ areas containing leukemic cells and various other populations, and ‘control’ areas in the non-leukemic BM (region types are illustrated in **Fig. 4A-C**; representative images of ROI selection are shown in **Fig. S5A**). In total, we successfully sequenced 143 regions with an average of 335 cells per region (range: 126 to 826 cells): 35 LA regions, 92 mixed regions, and 16 control regions. Our analysis revealed two main clusters of regions in the UMAP: cluster 1 contained both regions with LAs and mixed regions from biopsies with LAs, while cluster 2 consisted of mixed regions from biopsies without these aggregates, and control regions (**Fig. 4D**). Consequently, we divided mixed regions in two distinct groups: areas from biopsies with LAs (MIXED1, N=48), and those from biopsies without such aggregates (MIXED2, N=44). All three TLS-specific gene signatures were significantly enriched in LA regions in comparison to both mixed- and control regions (**Fig. 4E-F** and **S5B**). Thus, while the identified LAs lacked the typical organization of mature TLS, we did identify enrichment of the Tfh- and TLS imprint signatures representative of mature TLSs.

**Figure 4.**
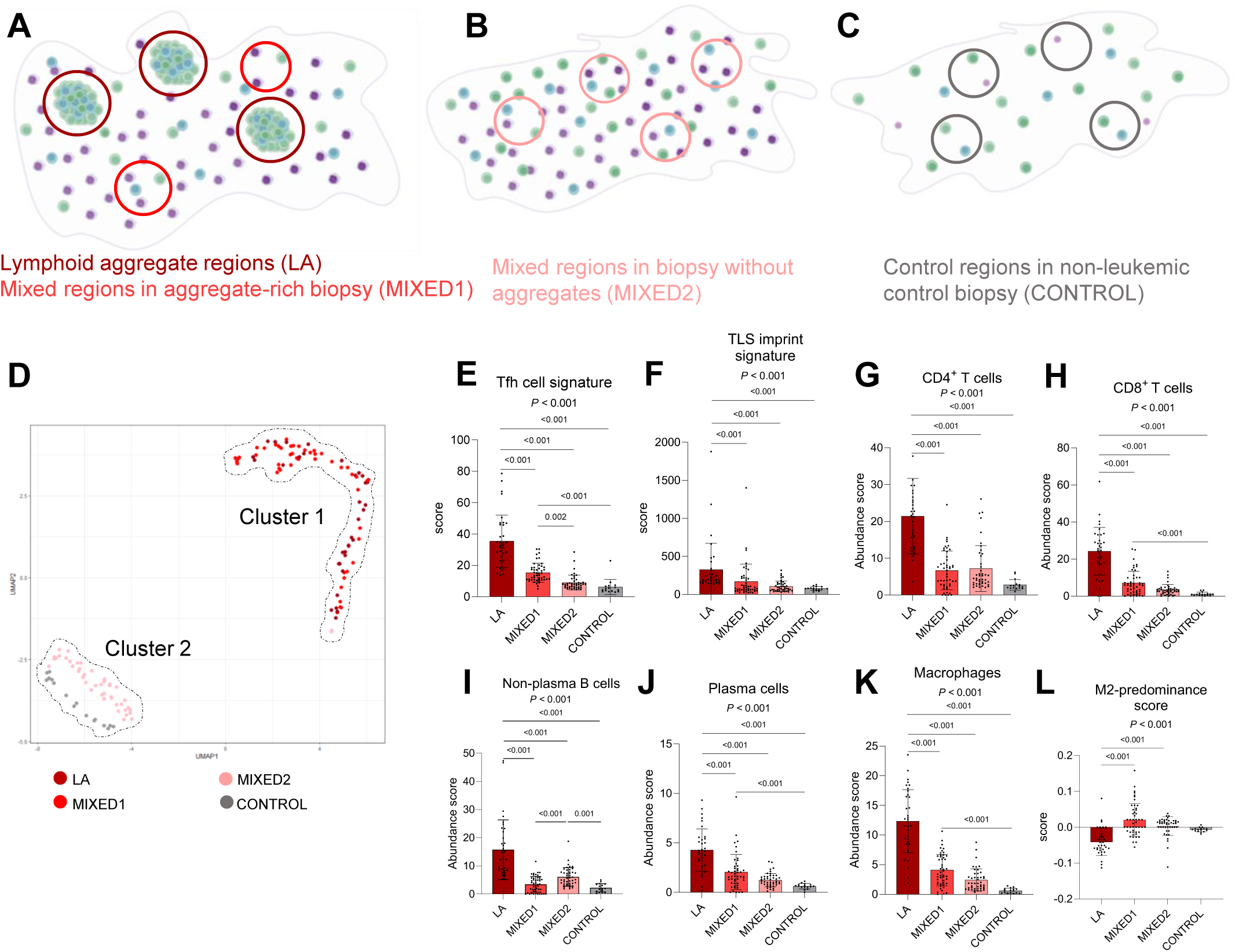
Composition of lymphoid aggregates in immune-infiltrated pediatric AML. (A-C) Overview of the different types of regions profiled using spatial transcriptomics in immune-infiltrated biopsies with lymphoid aggregates (A), immune-infiltrated biopsies without lymphoid aggregates (B), and non-leukemic biopsies (C). Green cells indicate T cells, blue cells indicate B cells, and pink/purple cells indicate AML blasts or normal myeloid cells. These examples do not necessarily reflect the actual abundance of these subsets. LA: lymphoid aggregate. (D) UMAP of transcriptomic profiles of various region types, organized into two separate clusters. (E-F) Comparison of the expression of the ‘Tfh’ (E), and ‘TLS imprint’ (F) signatures across different region types (Kruskal-Wallis followed by Dunn’s multiple comparisons test). (H-L) Deconvoluted abundance of various cell subsets and the M2-predominance score compared across different region types (Kruskal-Wallis followed by Dunn’s multiple comparisons test). In case of multiple p-values, the upper one is associated with the Kruskal-Wallis test, while the lower one(s) reflects the result of Dunn’s multiple comparison test.

To gain insight into differences in immune and stromal cell types between LA and other BM regions, we performed immune deconvolution by integrating spatial transcriptomic data with single-cell (sc) and flow-sorted bulk RNA-sequencing data of microenvironmental cell populations^51^. This analysis revealed that LA regions had an increased abundance of CD4^+^ T cells, CD8^+^ T cells, non-plasma B cells, plasma cells, and macrophages (**Fig. 4G-K)**. In both LA and MIXED1-regions, the CD4^+^:CD8^+^ T cell ratio was lower compared to MIXED2- and control regions (approximately 1:1 in both vs. 2:1 and 3:1, respectively; **Fig. S5C**), indicating a relatively higher abundance of CD8^+^ T cells in the former regions. When considering macrophage polarization, we noted a significantly lower M2-predominance score in LA regions compared to both mixed regions, consistent with the negative correlation between M2-predominance and BM T cell infiltration observed above (**Fig. 4L**; **Fig. 2F**). Moreover, endothelial cells and fibroblasts were enriched in LA regions compared to MIXED1-regions (**Fig. S5D-E**). Additionally, NK- and dendritic cells were distributed evenly across AML biopsies, while MIXED2 regions had a significantly higher abundance of neutrophils compared to both LA and MIXED1-regions (**Fig. S5F-H**). To confirm our deconvolution results, we performed differential gene expression analysis between LA, mixed, and control regions. In LA regions compared to non-aggregate regions within the same biopsies (MIXED1), we found upregulation of many T- and B cell genes (e.g., *CD3D, CD8B, CD79A*), as well as genes associated with TLS formation in solid cancers (*LTB, CCL19)* and B cell recruitment *(CXCL13*; **Fig. S6A**). Furthermore, immunoglobulin-related genes (*IGHG1*, *IGHG3*, *IGHG4*, *IGKC*) were significantly enriched in LA compared to MIXED1-regions, consistent with an increased abundance of plasma and/or memory B cells (**Fig. S6A**). Similar results were observed when comparing LA regions with MIXED2- and control regions (**Table S4**).

Given the combination of a lack of distinct T- and B cell zones and an increased estimated abundance of plasma cells in these LAs, we next sought to perform a more in-depth characterization of the B cells in these regions. To do so, we again applied immune deconvolution by integrating our spatial transcriptomic data with another scRNA-sequencing reference dataset (**Fig. 5A**). In this case, we used a comprehensive scRNA-sequencing dataset of pediatric tonsillar B cells^52^, which included naïve B cells, germinal center B cells, plasmablasts, and memory B cells, as a reference. In line with the lack of a follicle-like morphology of LAs, we did not identify germinal center B cells in these regions (**Fig. 5B**). Similarly, naïve B cells were absent. The most abundant B cells were memory B cells, while a smaller fraction consisted of plasmablasts, together indicating the presence of differentiated B cell subsets in these aggregates (**Fig. 5B**). Since LA regions showed increased expression of the cytotoxicity-related genes *GZMA* and *GZMK* compared to neighboring regions (**Fig. S6A**), we also aimed to investigate the cytotoxic capacity of CD8^+^ T cells in these regions. Accordingly, we performed immune deconvolution of our spatial transcriptomic dataset using a scRNA-sequencing dataset of BM CD8^+^ T cells from adult AML patients^16^. This reference dataset included naïve-, memory-, cytotoxic-(CTL), mucosal-associated invariant T-(MAIT), and ‘dysfunctional’ CD8^+^ T cells. The CD8^+^ T cells classified as dysfunctional were characterized by increased expression of multiple immune checkpoint receptor (IR) genes, such as *PDCD1*, *LAG3*, and *HAVCR2*, and reduced expression of cytotoxicity-related genes such as *GZMB*, *GNLY*, and *PRF1*. Notably, deconvolution revealed that these potentially dysfunctional CD8^+^ T cells were enriched in LA regions compared to mixed- and control regions, at the expense of both naïve- and cytotoxic CD8^+^ T cells (**Fig. 5C-D**). Moreover, memory CD8^+^ T cells were not identified in any of the biopsies. The increased proportion of potentially dysfunctional CD8^+^ T cells in LA regions prompted us to investigate whether these T cells were truly dysfunctional (i.e. no cytotoxic potential). Accordingly, we examined the expression of the cytotoxicity marker granzyme B (GZMB) in CD8^+^ T cells that expressed at least two immune checkpoint receptor markers (IR^++^/^+++^; PD-1, LAG3, and/or TIM-3) in a subset of the LA-rich biopsies that had been characterized using spatial transcriptomics (n=2 biopsies with six LAs in total). Using multiplex immunofluorescence with antibodies against CD3, CD8, PD-1, LAG3, TIM-3, and GZMB, in combination with standard IHC for CD20, we identified that 58% (range: 30 to 100%) of CD3^+^CD8^+^IR^++/+++^ T cells in LAs expressed GZMB (representative images shown in **Fig. 5E**; **Fig. 5F**), suggesting that more than half of the potentially dysfunctional CD8^+^ T cells still had cytotoxic capacity. In addition, since TLS-associated regulatory T cells (Tregs) have been found to attenuate the positive prognostic effect of TLS-associated CD8^+^ T cells in human non-small cell lung cancer, colorectal cancer, and soft tissue sarcoma^45, 53, 54^, we assessed the presence of putative Tregs (CD3^+^FOXP3^+^) in LAs in the same subset of biopsies (n=2). In the six examined LAs, we observed very few Tregs (<1%), suggesting that Tregs are hardly present in LAs in pediatric AML (representative image shown in **Fig. S6B; Fig. S6C**). Taken together, our integrative spatial analyses demonstrate that LAs in the BM of pediatric AML are specifically enriched for CD8^+^ T cells, memory B cells, plasma cells and/or plasmablasts, and M1-like macrophages. Despite the lack of germinal center B cells and separate T- and B cell zones, the presence of memory B cells, plasma cells and/or plasmablasts, and immunoglobulin gene expression suggests that LAs in pediatric AML are sites of B cell differentiation. Furthermore, the presence of CD8^+^ T cells expressing multiple immune checkpoint markers, yet not fully exhausted, within LAs, encourages future investigations into the potential of leveraging these LA-associated T cells for immunotherapeutic efficacy.

**Figure 5.**
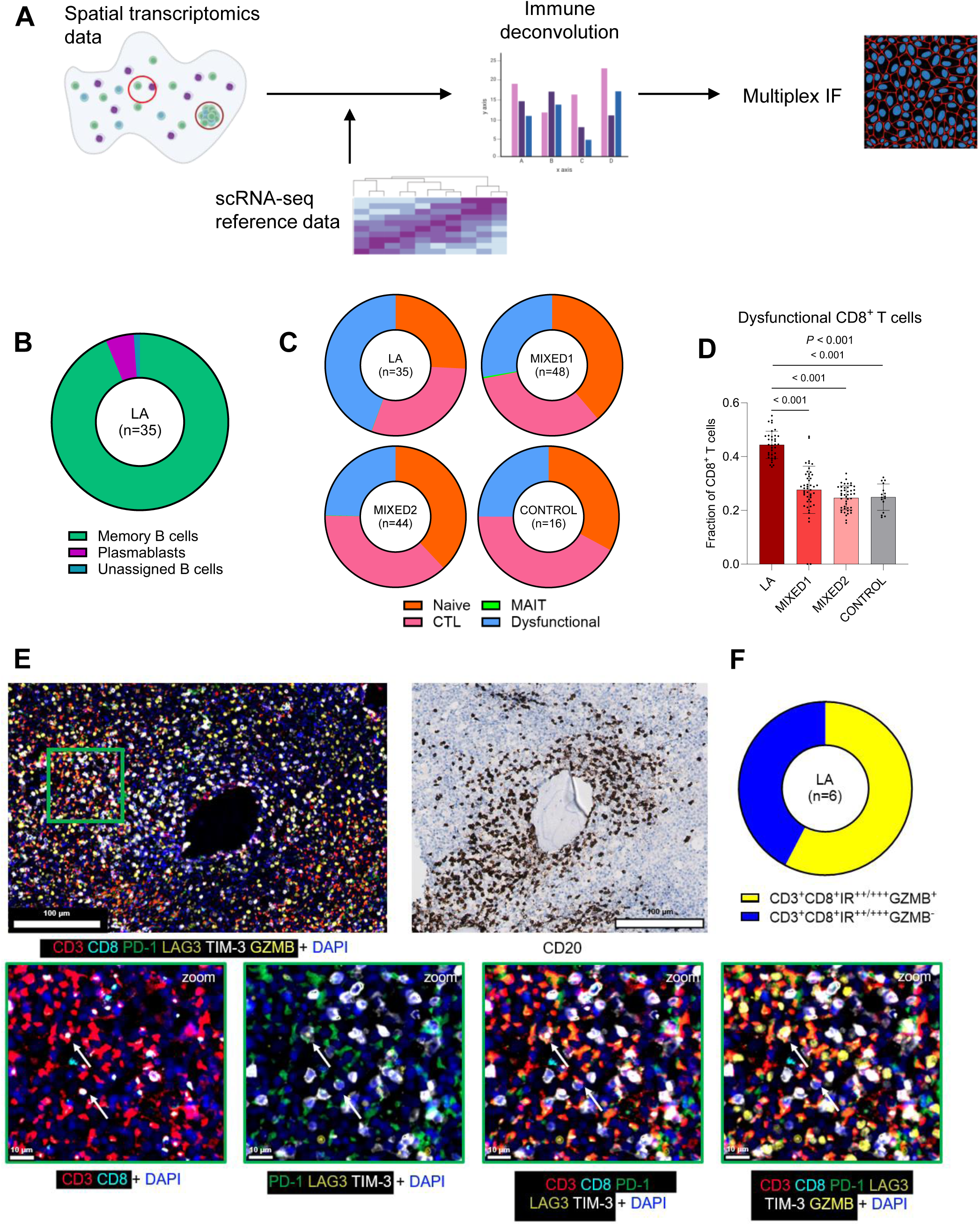
In-depth characterization of T- and B cells in lymphoid aggregates. (A) Schematic overview of the analysis approach applied to the spatial transcriptomics dataset and the subsequent multiplex immunofluorescence (IF). scRNA-seq: single-cell RNA-sequencing. (B) Proportions of memory B cells, plasmablasts, and unassigned B cells in lymphoid aggregates (LA). (C) Proportions of naïve-, cytotoxic (CTL), mucosal-associated invariant T (MAIT)-, and potentially dysfunctional CD8^+^ T cells in lymphoid aggregate, mixed, and control regions. (D) Comparison of the deconvoluted proportions of potentially dysfunctional CD8^+^ T cells in lymphoid aggregate, mixed, and control regions (Kruskal-Wallis followed by Dunn’s multiple comparisons test). In case of multiple p-values, the upper one is associated with the Kruskal-Wallis test, while the lower ones reflects the result of Dunn’s multiple comparison test. (E) Representative image of the multiplex immunofluorescence analysis of a lymphoid aggregate. The names below each image indicate which antibodies are shown. The green boxes on the lower row are zoomed in on the part of the biopsy in the green box in the upper left image. (F) The proportion of CD3^+^CD8^+^ T cells that expressed two or three inhibitor receptors (IR^++^/^+++^) positive (or not) for granzyme B (GZMB).

## Discussion

In this study, we performed a multidimensional assessment of the tumor immune microenvironment in pediatric AML. We demonstrated that nearly one-third of pediatric AML cases has an immune-infiltrated BM, which is characterized by a decreased M2-:M1-like macrophage ratio. Furthermore, we detected the presence of large T cell networks, both with and without colocalizing B cells, in the BM and dissected the cellular composition of T- and B cell-rich aggregates using spatial transcriptomics. These analyses revealed that these LAs are hotspots of CD8^+^ T cells, memory B cells, plasma cells and/or plasmablasts, and M1-like macrophages. Collectively, our study provides a multidimensional characterization of the BM immune microenvironment in pediatric AML and indicates starting points for further investigations into immunomodulatory mechanisms in this devastating disease.

Understanding the prevalence of distinct immune phenotypes and associated targetable immune-evasion mechanisms in the BM of pediatric AML is key for better prospective selection of patients in future immunotherapy trials^33, 45^. Although we identified a subset with high abundance of T cells, many pediatric AML cases had an immune-depleted BM microenvironment. We observed that *FLT3*-ITD and/or *NPM1* mutations were associated with low T cell infiltration in the BM of pediatric AML, consistent with data in adult AML^33^. Although we did not investigate the direct consequences of these molecular alterations, such as alterations in the expression of immunomodulatory surface molecules^38, 39^, we identified M2-like macrophage predominance in immune-depleted cases. The observed negative correlation between the M2-:M1-like macrophage ratio and the extent of T cell infiltration is consistent with data from (preclinical) studies in other cancers, suggesting a role for M2-like macrophages (or a lack of M1-like macrophages) in limiting T cell infiltration to the leukemic BM^40, 41^. Consequently, targeting M2-like macrophages (e.g., using checkpoint blockade) to overcome their immunosuppressive functions could be an attractive therapeutic strategy for immune-depleted AML^55, 56^.

In addition, our integrative spatial analyses showed that T cells in the BM can be organized into multicellular aggregates. Although T- and B cell-rich LAs did not contain distinct T- and B cell zones and germinal center B cells typically seen in mature TLSs in solid cancers, the localized enrichment of memory B cells, plasma cells and/or plasmablasts, and immunoglobulin gene expression suggests that these aggregates are sites of B cell maturation. Moreover, the 1:1 ratio of CD4^+^ and CD8^+^ T cells observed in LAs has also been associated with mature TLSs^57^. Notably, CD8^+^ T cells in LAs were found to be in different cell states, including a subset that expressed multiple immune checkpoint markers. GZMB-expression was identified in more than half of these cells, suggesting that these CD8^+^ T cells are not fully exhausted and may be susceptible to ICI therapy^58^. In addition to T- and B cell-rich immune aggregates, we detected large T cell networks with sparse B cells. Since these networks have also been associated with ICI response in solid cancers, further investigations into the different types of immune aggregates in AML and their relevance for immunotherapy response are needed^43, 46, 59, 60^. For instance, do large T cell-dominant networks relate to LAs, and do they (differentially) associate with ICI efficacy in AML? Since TLS are considered to be sites where tumor-specific T- and B cell immune responses may be generated^23–25^, future research also needs to investigate whether immune aggregates in AML are tumor-directed.

Altogether, our analyses deepen the understanding of the BM immune microenvironment in AML and provides an impetus to explore how intratumoral immune aggregates could be exploited for improving immunotherapy outcomes. Further, our work provides a framework for leveraging spatial transcriptomics to interrogate the spatial organization of the leukemic BM. Advances in spatial transcriptomic techniques now allow for investigating the spatial dimension of hematological malignancies at subcellular resolution, opening an exciting path towards new discoveries in the field of AML^61, 62^.

## Acknowledgements

We would like to thank all patients and/or their families for their generous consent for the research use of these samples; the staff of the University Medical Center Utrecht Tissue Facility for their excellent immunohistochemistry service (Domenico Castigliego, Petra van der Weide, Petra van der Kraak, Erica Siera, Karina Timmer, Sven van Kempen), and the team at Utrecht Sequencing Facility for performing the NanoString experiments (nCounter and GeoMx) and for providing assistance with data-analysis (dr. Ies Nijman, Robin Geene, Pim Kloosterman); dr. Thierry van den Bosch, dr. Ravian van Ineveld, Ella de Boed, Nienke van Herk, Thijs van den Broek, Maurice de Haan, and Susanne Gamas Vis provided help with imaging experiments and/or analysis; Dr. Matthew S. Davids for clinical data of patients treated on ETCTN/CTEP 9204; Dr. Ivette Deckers and dr. Annette Gijsbers (PALGA) performed essential work for the acquisition of bone biopsies from other hospitals; prof. dr. Gertjan Kaspers and dr. Bianca Goemans aided with identifying potential patients, the biobank staff (Jantien Woudstra, Marion Koopmans, dr. Edwin Sonneveld) helped to identify patient material, and Arie Maat aided with the sectioning of bone biopsy sections; dr. Caroline Lindemans, members from the Heidenreich group (dr. Katarzyna Szoltysek, dr. Farnaz Barneh, dr. Mauricio Ferrao-Blanco, Elizabeth Schweighart, Nina van der Wilt), the Van Heesch group (dr. Ana Pinheiro-Lopes), and the single-cell sequencing facility (dr. Lindy Visser) at the Princess Máxima Center for Pediatric Oncology for carefully reading the manuscript and/or fruitful discussions. Figures have been created using BioRender.com.

## Author contributions

Conceptualization: J.B.K, I.W., M.A.V., S.N., M.B., C.M.Z., O.H. Methodology: J.B.K, I.W., M.A.V., M.F., S.N., M.B., C.M.Z., O.H. Data acquisition: J.B.K., M.A.V., F.B., J.I.M.-E. H.G.-K., R.M, K.B.C., B.P., K.P., H.H. Data analysis and interpretation: J.B.K., I.W., L.P., F.B., A.P., J.I.M.-E., J.S.G., S.J.R., C.J.W., H.H. Writing – Original Draft: J.B.K.; Writing – Reviewing & Editing: all authors; Supervision: S.N., M.B., C.M.Z., O.H.

## Competing interests

J.S.G. reports serving on steering committee and receiving personal fees from AbbVie, Genentech, and Servier and institutional research funds from AbbVie, Genentech, Pfizer, and AstraZeneca.

S.R. receives research support from Bristol-Myers-Squibb and KITE/Gilead, and is a member of the SAB of Immunitas Therapeutics.

C.J.W. holds equity in BioNTech, Inc and receives research support from Pharmacyclics.

C.M.Z. receives institutional research support from Pfizer, Abbvie, Takeda, Jazz, Kura, Gilead, and Daiichi Sankyo, provides consultancy services for Kura, Bristol-Myers-Squibb, Novartis, Gilead, Incyte, and Syndax, and serves on advisory committees for Novartis, Sanofi, and Incyte.

O.H. receives institutional research support from Syndax and Roche.

The remaining authors declare no competing financial interests.

## Data availability

Sequencing data can be accessed from the Gene Expression Omnibus (nCounter data: GSEXXX; GeoMx data: GSEXXX; both normalized counts [GSE IDs will be available upon approval]) and from the European Genome-phenome Archive (EGA) database (https://ega-archive.org/studies/EGAXXX; raw nCounter and GeoMx data [idem]).

## Funding

This work has been funded in part by a KIKA (329) program grant to O.H. L.P. is a Scholar of the American Society of Hematology, participant in the BIH Charité Digital Clinician Scientist Program funded by the DFG, the Charité – Universitätsmedizin Berlin, and the Berlin Institute of Health at Charité (BIH) and is supported by the Max-Eder program of the German Cancer Aid. J.S.G. is supported by the Conquer Cancer Foundation Career Development Award, Leukemia and Lymphoma Society Translational Research Program Award, and NIH K08CA245209. NCI CTEP provided study drug (Ipilimumab) support.

This work was supported by National Institutes of Health, National Cancer Institute grant P01CA229092 (C.J.W.), UM1CA186709 (Principal Investigator: Geoffrey Shapiro), National Cancer Institute Cancer Therapy Evaluation Program, Bristol-Myers Squibb, and LLS Therapy Accelerator Program. This work was further supported by the CIMAC-CIDC Network. Scientific and financial support for the CIMAC-CIDC Network is provided through National Institutes of Health, National Cancer Institute Cooperative Agreements U24CA224319 (to the Icahn School of Medicine at Mount Sinai CIMAC), U24CA224331 (to the Dana-Farber Cancer Institute CIMAC), U24CA224285 (to the MD Anderson Cancer Center CIMAC), U24CA224309 (to the Stanford University CIMAC), and U24CA224316 (to the CIDC at Dana-Farber Cancer Institute). The CIMAC-CIDC website is found at https://cimac-network.org/.

## Table Legends (tables are provided in a separate excel file)

**Supplementary Table 1. Clinical characteristics of the primary study cohort, supplemented with information on immune-infiltration, performed assays, and T cell networks.**

**Supplementary Table 2. List of significantly up- or downregulated pathways comparing immune-infiltrated versus immune-depleted cases using ssGSEA.**

**Supplementary Table 3. Clinical characteristics of the adult AML cohort treated with ipilimumab-based therapy.**

**Supplementary Table 4. Differentially expressed genes (DEGs) between regions in the spatial transcriptomics (tx) dataset identified by linear mixed-effect modelling (LMM).**

**Supplementary Table 5. Target antigens, antibody clones and suppliers of antibodies used for pediatric AML bone marrow IHC/multiplex immunofluorescence.**

**Supplementary Table 6. Target antigens, antibody clones, suppliers, dilution of markers, diluents, and antigen retrieval conditions used for adult AML bone marrow multiplex immunofluorescence.**

## Supplemental information titles and legends

**Supplementary Figure 1.**
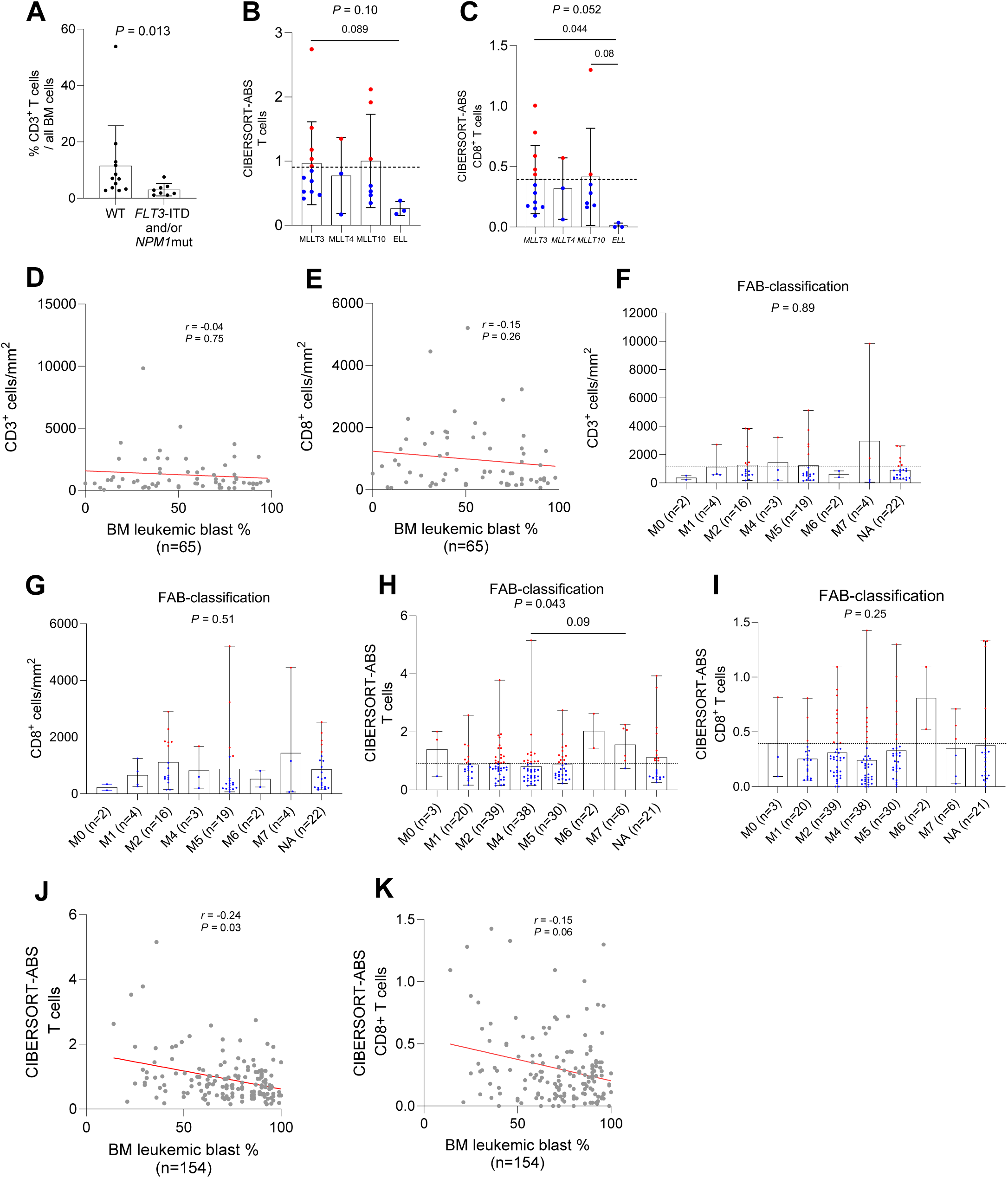
AML blasts and differentiation stage in relation to T- and CD8^+^ T cell infiltration in the bone marrow. (A) Comparison of the percentage of CD3^+^ T cells out of all bone marrow (BM) cells between diagnostic pediatric AML cases with normal karyotype and no identified molecular aberrations (wildtype) and those with a *FTL3*-ITD and/or *NPM1* mutation (*NPM1*mut; Mann-Whitney test). The T cell proportions were retrieved from diagnostic flow cytometry reports. These patients are part of an independent cohort, not the primary study cohort. (B-C) Comparison of the CIBERSORTx-based estimated absolute (ABS) abundance of T-(B) and CD8^+^ T cells (C) between pediatric AML patients (TARGET-AML cohort) with different *KMT2A*-rearrangements. The names on the x-axis indicate the *KMT2A*-fusion partners. The dashed lines indicate the median estimated abundance of T- and CD8^+^ T cells in four non-leukemic controls. Examined using the Kruskal-Wallis test followed by Dunn’s multiple comparisons test. In case of multiple p-values, the upper one is associated with the Kruskal-Wallis test, while the lower one(s) reflects the result of Dunn’s multiple comparison test. (D-E) Correlation plot between the normalized number of CD3^+^ (D) and CD8^+^ (E) T cells and leukemic blasts in the bone marrow (blast % available for 65 cases), calculated using Spearman correlation. (F-G) Comparison of the normalized abundance of CD3^+^ (F) and CD8^+^ (G) T cells in the bone marrow across AML differentiation stages (FAB-classifications; Kruskal-Wallis followed by Dunn’s multiple comparisons test). Data are presented as mean plus range. The dashed lines indicate the median T- and CD8^+^ T cell abundance in non-leukemic controls. (H-I) Comparison of the estimated abundance of T-(H) and CD8^+^ (I) T cells in the bone marrow of TARGET-AML cases across AML differentiation stages (FAB-classifications; Kruskal-Wallis followed by Dunn’s multiple comparisons test). Data are presented as mean plus range. The dashed lines indicate the estimated median T- and CD8^+^ T cell abundance in non-leukemic controls. (J-K) Correlation plot between the estimated number of T-(J) and CD8^+^ (K) T cells and leukemic blasts in the bone marrow (blast % available for 154 cases) of TARGET-AML cases, calculated using Spearman correlation.

**Supplementary Figure 2.**
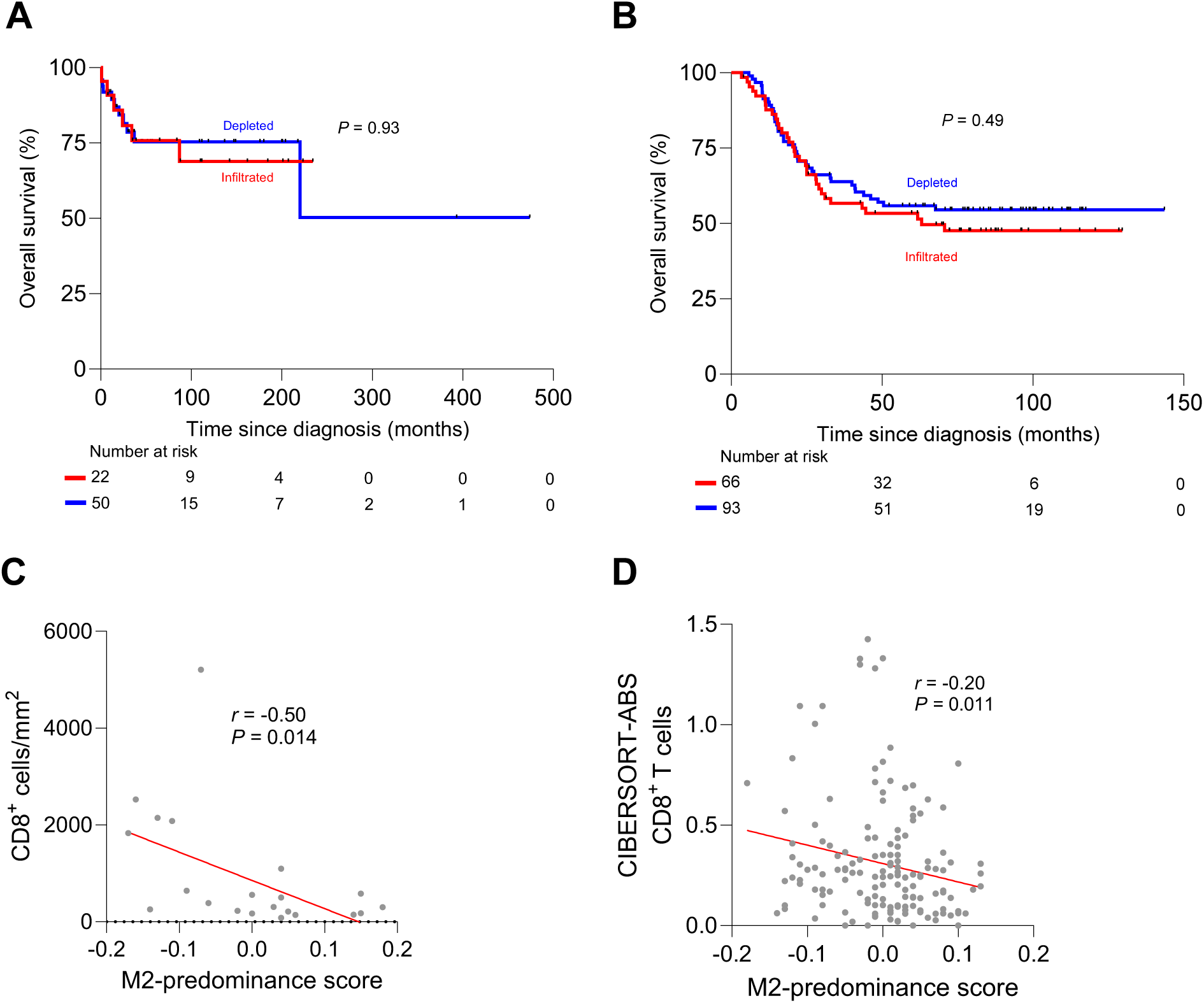
Overall survival and M2-like macrophage abundance in immune-infiltrated and immune-depleted cases across two cohorts. (A-B) Overall survival between immune-infiltrated and immune-depleted pediatric AML cases in our primary study cohort (A) and in the TARGET-AML cohort (B). (C-D) Correlation plots of the correlation between the M2-predominance score and the normalized (C; our cohort) or estimated (D; TARGET-AML cohort) abundance of CD8^+^ T cells, calculated using Spearman correlation.

**Supplementary Figure 3.**
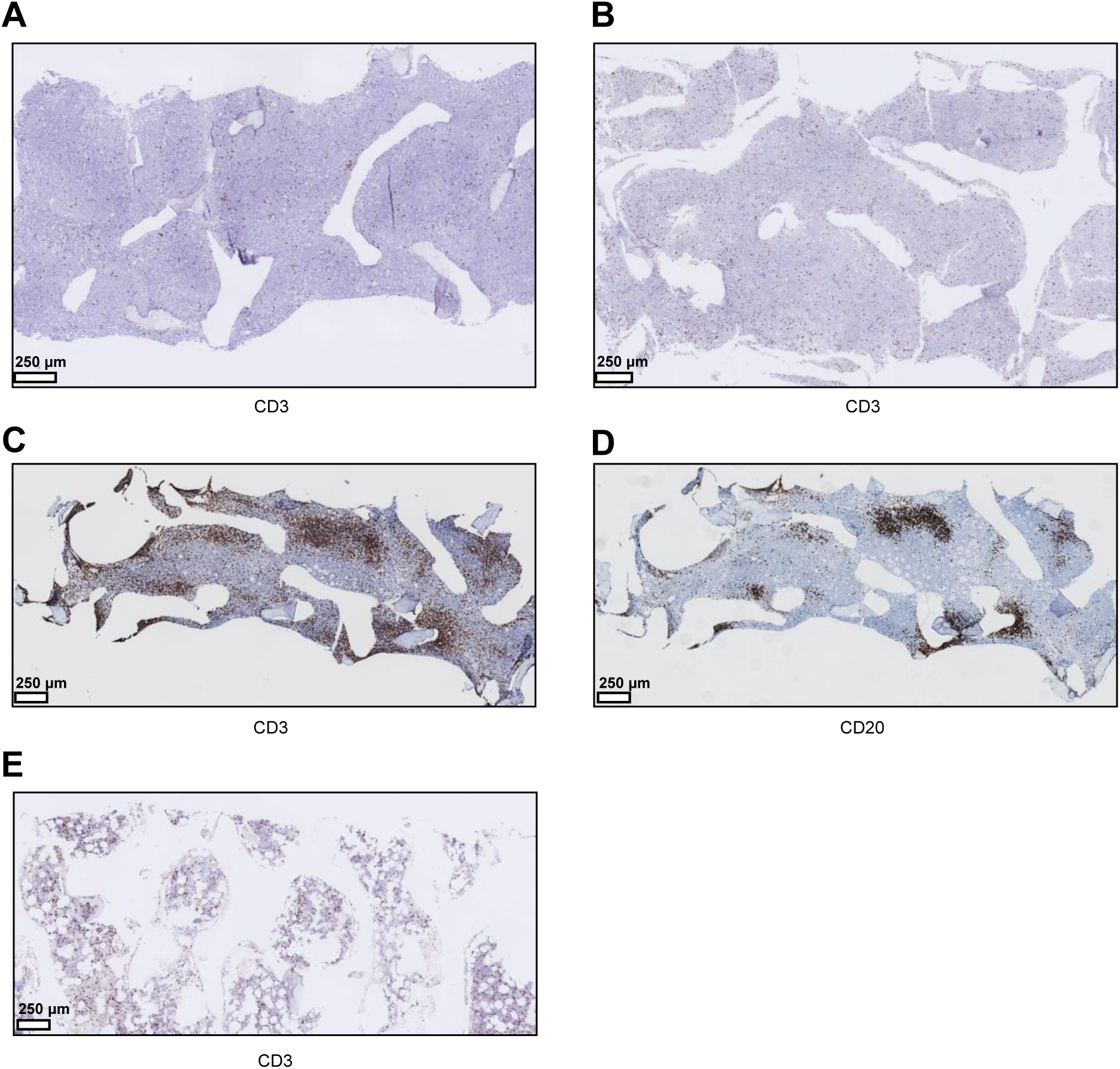
Representative images of bone marrow biopsies of immune-infiltrated and immune-depleted pediatric AML, and a non-leukemic control. (A-B) Representative images of CD3^+^ T cells in the bone marrow of immune-depleted cases. (C-D) Representative images of large T cell networks (C) that colocalized with a dense network of B cells (lymphoid aggregates) (D). (E) Representative image of CD3^+^ T cell infiltration in the bone marrow of a non-leukemic control.

**Supplementary Figure 4.**
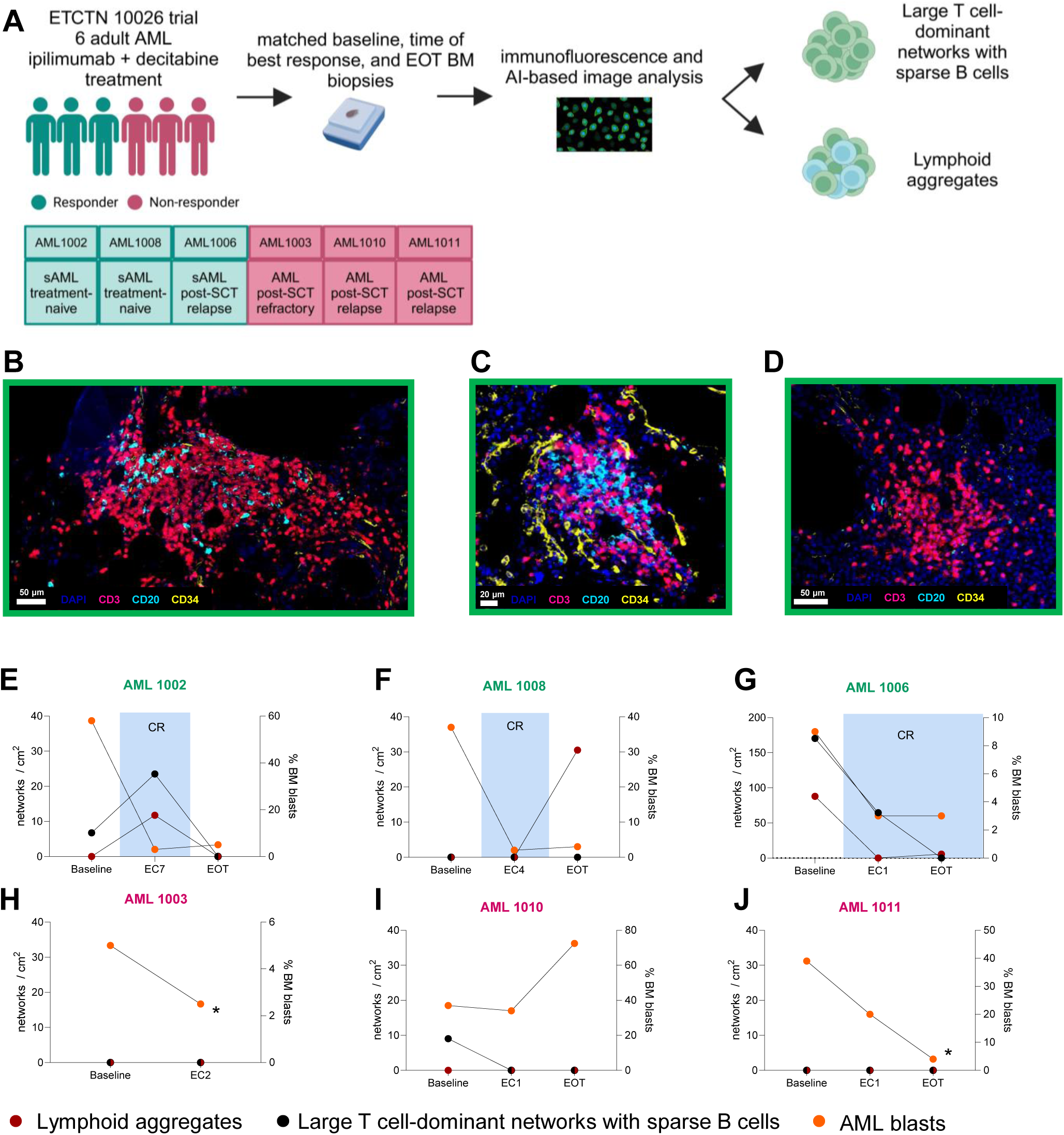
Immune aggregates in the bone marrow of adult AML cases treated with ipilimumab-based treatment. (A) Schematic overview of adult AML cases that were treated with ipilimumab and decitabine, the used techniques, and the examined variables. The table indicates the patient IDs, whether patients responded (yes=green, purple=no) to these therapies, whether the patients had primary AML (AML) or secondary AML (sAML), and whether they were treated in the treatment-naïve or post-stem cell transplantation (SCT) setting. (B-D) Representative multiplex immunofluorescence images of lymphoid aggregates (B-C) and large T cell-dominant networks with sparse B cells (D). (E-J) Longitudinal overview of lymphoid aggregates, T cell-dominant networks, and AML blasts in bone marrow biopsies before treatment according to the clinical trial (baseline), at time of best response, or at end of treatment (EOT). Presented blast percentages are the average of the aspirate and biopsy blast percentages. EC=end of course; the number (e.g., EC1) indicates which course was given (e.g., EC1 indicates that the biopsy was taken after course 1). Green IDs indicate responders, purple IDs indicate non-responders. The AML blast percentage was the average of the percentages in the aspirate and the core biopsy. CR=complete morphologic remission with or without complete count recovery and no signs of leukemia elsewhere. *AML1003 had an aplastic bone marrow without hematologic recovery, and AML1011 had 5% blasts by histology.

**Supplementary Figure 5.**
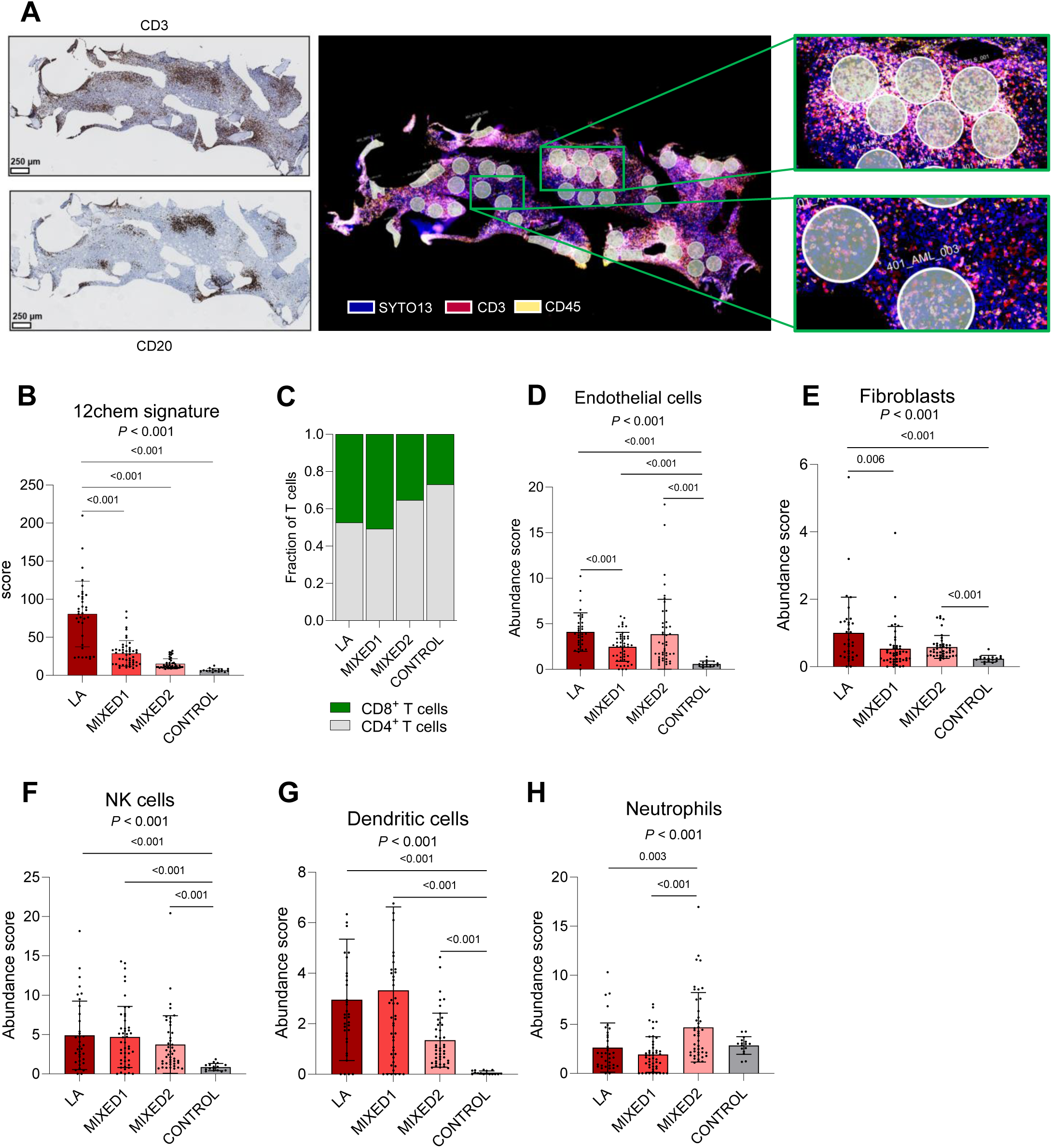
Composition of lymphoid aggregates in immune-infiltrated pediatric AML. (A) Representative images of the region of interest (ROI) selection on the GeoMx Digital Spatial Profiling platform. On the left, both the CD3 (T cell) and CD20 (B cell) stains are shown (DAB, both in brown). On the right, magnifications of immune-aggregate regions (above) and mixed regions (below) are shown. Selection of ROIs was further aided by overlaying stains of CD34 and CD117 (Supplementary Methods). (B) Comparison of the expression of the ‘12chem’ signatures across different region types (Kruskal-Wallis followed by Dunn’s multiple comparisons test). (C) Proportions of CD4^+^ and CD8^+^ T cells in lymphoid aggregate (LA), mixed, and control regions. (D-H) Deconvoluted absolute abundance of various cell subsets across several region types (Kruskal-Wallis followed by Dunn’s multiple comparisons test). In case of multiple p-values, the upper one is associated with the Kruskal-Wallis test, while the lower one(s) reflects the result of Dunn’s multiple comparison test.

**Supplementary Figure 6.**
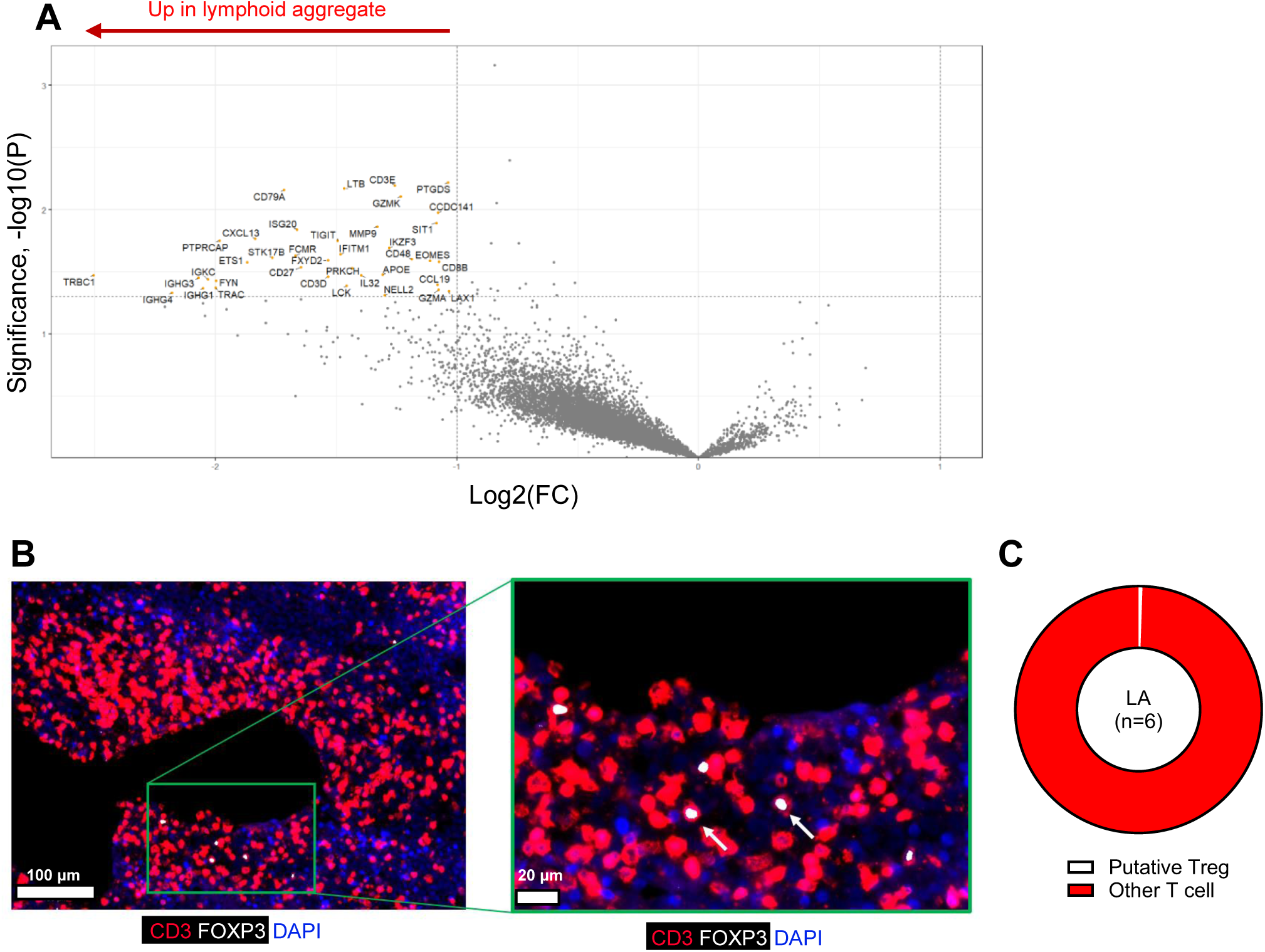
Upregulated genes and the presence of regulatory T cells in lymphoid aggregates in immune-infiltrated pediatric AML. (A) Volcano plot of genes differentially expressed in lymphoid aggregates compared to MIXED1 regions, generated using linear mixed-effect modelling. FC: fold change. (B) Representative images of putative Tregs (CD3^+^FOXP3^+^) in lymphoid aggregates (CD3^+^ T cells). The image on the right reflects a zoom of the region in the green box in the left image. The names below the images reflect the antibodies and the associated colors in the images. (C) Proportion of putative Tregs among all CD3^+^ T cells in lymphoid aggregates (LA).

## Notes

### Author Declarations

This study complies with all relevant ethical regulations and was approved by the Institutional Review Boards of the Princess Maxima Center for Pediatric Oncology (PMCLAB2021.207 & PMCLAB2021.238), the Scientific Committee of the Dutch Nationwide Pathology Databank (PALGA: lzv2021-82)64 and at participating sites of the ETCTN/CTEP 10026 study after written informed consent was obtained. All patients treated at the Princess Maxima Center, Aarhus University Hospital, and Dana-Farber Cancer Institute provided written consent for banking and research use of these specimens, according to the Declaration of Helsinki. BM biopsy tissues acquired from external biobanks (n=28) were leftover material from standard care procedures and therefore, no informed consent was acquired, according to Dutch legislation and the code of conduct of the Committee on Regulation of Health Research (COREON).

### Summary of Updates

-We revisited the approach to analyzing the association between AML-associated genetic alterations and T cell infiltration in the BM, and identified a link between FLT3-ITD and/or NPM1 mutations and low T cell infiltration. This association was confirmed in an independent cohort of 20 pediatric AML patients. -Additionally, we integrated two comprehensive single-cell RNA-sequencing datasets with our spatial transcriptomics data to further investigate the B- and CD8+ T cell composition of lymphoid aggregates. -We also utilized multiplex immunofluorescence to examine the cytotoxic capacity of potentially dysfunctional CD8223 cl:114 T cells in lymphoid aggregates, as well as the presence of regulatory T cells. -Furthermore, we have added illustrations to each main figure to depict the workflow of our analyses and have been more succinct in referring to specific agents and terminology.

