## Supplemental Data for "A multidimensional analysis reveals distinct immune phenotypes and the composition of immune aggregates in pediatric acute myeloid leukemia"

to

Koedijk et al.

**Supplementary Methods**

**Ethical regulation**

Pediatric AML bone marrow (BM) biopsy tissues acquired from external biobanks (n=28) were leftover material from standard care procedures and therefore, no informed consent was acquired, according to Dutch legislation and the code of conduct of the Committee on Regulation of Health Research (COREON).

**Human patient samples**

As non-leukemic controls, FFPE BM biopsies from age- and sex-matched children with treatment-naïve early-stage rhabdomyosarcoma were obtained from the Princess Máxima Center Biobank (n=10). An experienced hemato-onco pathologist confirmed that these control biopsies resembled normal hematopoiesis and that there was no malignancy infiltrating the BM. AML patients were all treated using intensive anthracycline- and cytarabine-based chemotherapy and allogeneic stem cell transplantation in selected high-risk cases^1^.

**Response definitions of ETCTN 10026 clinical trial in adult AML patients**

We used the definitions of response according to the 2003 International Working Group criteria for acute myeloid leukemia^2^. All responders achieved a morphologic CR and AML1010 had >5% blasts. Although AML1011 had <5% blasts in the aspirate, histology indicated 5% blasts. Furthermore, AML1003 had <5% blasts but also an aplastic marrow without hematologic recovery and thus classified as a morphologic leukemia-free state (MLFS; part of the non-response categories).

**Immunohistochemistry/immunofluorescence and digital image analysis**

BM pediatric and adult AML cases were cut into consecutive sections of 4 and 5 μm, respectively. Conventional IHC was performed for CD3 (T cells), CD3-CD4 (duplex; for CD4^+^ T cells and CD4^+^ AML cells), CD8 (mainly CD8^+^ T cells but may also stain rare CD8^+^ NK- and dendritic cells), CD20 (B cells), CD34, CD117, and CD15 (immunophenotype-based AML markers) on pediatric AML BM biopsies using a Ventana Benchmark Ultra (Roche, Basel, Switzerland) automated staining instrument according to manufacturers’ recommendations. A list of antibodies and suppliers is available in **Table S5**. Digital scans of stained slides were obtained using a NanoZoomer scanner (Hamamatsu, Shizuoka, Japan).

Multiplex immunofluorescence using DAPI and antibodies against CD3 (T cells), CD8 (CD8^+^ T cells), granzyme B (GZMB; cytotoxicity marker), PD-1, TIM-3, and LAG3 (all immune checkpoint receptors) was performed on two treatment-naïve pediatric AML biopsies (AML2 + AML6) using SignalStar^TM^ from Cell Signaling Technology as previously described^3^. Briefly, CD3, CD8, granzyme B, PD-1, TIM-3, and LAG were conjugated to oligonucleotides (oligos) and then validated in the SignalStar multiplex assay to assess the (CD8^+^) T cell compartment of the tumor microenvironment (a list of antibodies and suppliers is listed in **Table S5**). All primary antibodies were applied at once in one primary incubation step. Complimentary oligos with fluorescent dyes (channels: 488, 594, 647, and 750 nm) subsequently amplified the signal of up to four oligo-conjugated antibodies in the first round of imaging. Then, the first round of oligos and fluorophores were gently removed and a second batch of complementary oligos with fluorescent dyes was added to again amplify the signal of up to four additional oligo-conjugated antibodies. Staining of the biopsies was done on a BOND RX fully automated stainer (Leica Biosystems, Nußloch, Germany) after slide baking for 30 minutes at 60°C. Imaging was performed using a Leica DMi8 inverted microscope (Leica Camera AG, Wetzlar, Germany), including automated stage and 3D Thunder deconvolution, using a 20x dry objective equipped with a Leica DFC700GT camera. Slides were scanned and images were deconvoluted and stitched using LASX (Leica). Subsequently, images were analyzed in QuPath^4^ as described below. Multiplex immunofluorescence using DAPI and antibodies against CD3 (T cells) and FOXP3 (putative regulatory T cell marker) was performed on the same treatment-naïve pediatric AML biopsies (AML2 + AML6; consecutive slides) using the Ventana Benchmark Discovery (Ventana Medical Systems Inc; a list of antibodies and suppliers is provided in **Table S5**) and scanned (ZEISS Axioscan 7; Zeiss, Oberkochen, Germany) as previously described^5^.

Multiplex immunofluorescence using DAPI and antibodies against CD3 (T cells), CD20 (B cells), and CD34 (AML blasts and endothelial cells) was performed on baseline, time of best response, and end of treatment (EOT) BM biopsies of adult AML cases treated with ipilimumab-based therapy on the 10026 studies and imaged as previously described^2^. The time points analyzed for each patient were based on the availability of BM biopsies**.** One biopsy was excluded because of poor quality [EOT case AML1003]. Staining of the biopsies was completed on a BOND RX fully automated stainer (Leica Biosystems, Nußloch, Germany) after baking tissue sections for 3 hours at 60°C. The BOND RX then performed deparaffinization and rehydration with series of graded ethanol to deionized water. Subsequently, antigen retrieval was performed utilizing Epitope Retrieval Solution 1 (pH 6) or 2 (pH 9), as shown in **Table S6** (ER1, ER2, Leica Biosystems, Cat. AR9961, AR9640). The secondary antibodies that were used are described in **Table S6** as well. The antibody complex was incubated for 10 minutes with its corresponding Opal Fluorophore Reagent (Akoya Biosciences, Marlborough, MA, USA) for signal visualization. After the final fluorophore application, the samples were incubated in Spectral DAPI solution (Akoya) for 10 minutes. Lastly, samples were removed from the BOND, air-dried, and mounted using Prolong Diamond Anti-fade mounting medium (Life Technologies, Cat. P3695). Whole-slide images have been obtained using the PhenoImager HT multispectral imaging platform (Akoya), were spectrally unmixed (Inform 2.6, Akoya), and 20x regions were stitched together (QuPath, v0.3.2.)^3^. Immunofluorescence data with DAPI and CD3 performed on extramedullary biopsies was acquired from previously published work^4^. For all imaged slides, whole-slide digital image analysis was performed in QuPath^3^. Inside QuPath, the deep learning-based cell segmentation tool StarDist and the machine learning-based Random Trees classifier were used to quantify the number of positive cells per mm^2^ ^4, 6^. T cell proximity was established using Delaunay Triangulation^7^.

**Immune-related gene expression profiling**

Four consecutive sections of 10 μm from six immune-infiltrated and seventeen immune-depleted FFPE BM biopsies from a cytogenetically representative cohort of pediatric AML cases (patient characteristics in **Table S1**) were used to isolate RNA and to perform immune-related gene expression profiling with the 770-gene PanCancer IO 360 panel, as previously described (NanoString, Seattle, WA, USA)^8, 9^. After passing quality control, raw data were normalized using ROSALIND® according to NanoString’s recommendations (<https://rosalind.bio/>; San Diego, CA, USA). Normalized data were uploaded to the online iDEP platform and differentially expressed genes between immune-infiltrated and immune-depleted biopsies were identified using *DEseq2* with a false-discovery rate (FDR) cut-off of 0.05 and a minimum fold change of 2 (integrative Differential Expression and Pathway analysis; <http://bioinformatics.sdstate.edu/idep96/>; V.0.96)^10^. Pathway analysis was performed using single-sample gene set enrichment analysis with the GO Biological Processes and WikiPathways gene sets with an FDR cut-off of 0.05^11-13^. The M2-predominance score was estimated using a published gene signature of M2-like macrophages compared to M1-like macrophages, inside the TIDE environment (tide.dfci.harvard.edu/)^14, 15^.

**GeoMx Digital Spatial Profiling**

5 μm thick FFPE BM biopsy sections from six pediatric AML cases with an immune-infiltrated BM and two non-leukemic controls were put on three different slides and prepared for GeoMx Digital Spatial Profiling (DSP; NanoString), as previously described^16^. Slides were simultaneously incubated with immunofluorescent antibodies and GeoMx Whole Transcriptome Atlas profiling reagents. SYTO13 (S7575, Thermo Fisher,) was used for identification of nuclei, CD45 (NBP2-34528, Novus) for leukocytes, and CD3 (NBP2-54392AF647, Novus) for T cells. Stained slides were loaded onto the GeoMx instrument and scanned. ROIs were selected using the above-mentioned antibodies in combination with overlayed images of CD20, CD34, CD3-CD4 (duplex), and CD117 (IHC). Then, UV-photocleaved oligonucleotides were collected in separate wells and sequenced on the Nextseq2000 (Illumina, San Diego, CA, USA).

Raw data were normalized using Quartile 3 count (Q3) normalization in R (V.4.2.1) as per NanoString’s recommendations (code is available in the vignette of the *Geomxtools* package: https://bioconductor.org/packages/release/workflows/vignettes/GeoMxWorkflows/inst/doc/GeomxTools_RNA-NGS_Analysis.html). Batch correction was performed using Combat-seq^17^. Spatial Deconvolution was performed in the GeoMx DSP Analysis Suite using the safeTME reference (*SpatialDecon* package; cell reference profiles are available via <https://github.com/Nanostring-Biostats/CellProfileLibrary/blob/archive/safeTME-for-tumor-immune.csv>). Furthermore, we retrieved single-cell RNA-sequencing data from pediatric tonsillar B cells^18^ (sample BC005 was chosen since it had the highest number of cells, as done previously^19^) and adult AML bone marrow CD8^+^ T cells^20^ (all patients), and converted these two additional reference profiles using R (V4.2.1). Deconvoluted abundance scores were normalized for ROI-size and, in case of immune aggregates, for the ROI-area covered by these aggregates. The ‘12chem’, ‘Tfh’, and ‘TLS imprint’, and M2-predominance signatures were applied to Q3-normalized data and further normalized as mentioned above^19, 21-23^.

**External pediatric and adult AML datasets**

Bulk RNA-seq data (counts and transcripts-per-million; TPM) and matching clinical data of 159 treatment-naïve *de novo* pediatric AML cases generated as part of the TARGET-AML project was acquired via https://www.cBioportal.org^24, 25^. TPM values were used to estimate the abundance of total T- and CD8^+^ T cells using CIBERSORTx, as described previously (LM22 reference profile; https://cibersortx.stanford.edu/)^26-28^.

**Statistical analysis**

In case of no Gaussian distribution, differences between two independent groups and two paired groups were compared using the Mann-Whitney test and the Wilcoxon paired signed-rank test, respectively. For Gaussian-distributed data, unpaired and paired t-tests, respectively, were used. The correlation between two variables was evaluated using Spearman’s *r*. To measure the area under the receiver operating characteristic curve, we employed the Wilson-Brown method. To estimate the survival from diagnosis, the Kaplan-Meier’s methodology was employed. To assess the difference between survival estimates in different groups, the log rank test was used. In case of multiple comparisons and no Gaussian distribution of residuals, we employed the Kruskal-Wallis test followed by Dunn’s test for multiple comparisons including a Bonferroni correction. In case two p-values are shown, the upper one is associated with the Kruskal-Wallis test, while the lower one reflects the result of Dunn’s multiple comparison test.
